# Longitudinal epigenetic rewiring in lung immune cells in patients with post-COVID-19 condition

**DOI:** 10.1101/2025.02.06.25321750

**Authors:** Frida Nikesjö, Jelena Smiljanic, Shumaila Sayyab, David Martínez-Enguita, Mika Gustafsson, Martin Rosvall, Kristofer Hedman, Maria Lerm

## Abstract

**Background:** Post-COVID-19 condition (PCC) affects millions globally, presenting as persistent multisystem symptoms. Despite various hypotheses, the biological mechanisms underlying PCC remain unclear. Previous studies have linked PCC to alterations in the DNA methylome of blood immune cells, but the effects on lung cells over time remain unknown.

**Methods:** Patients (n=13) with persistent symptoms following COVID-19 in 2020-2021 donated blood and sputum samples at inclusion and after one year. Symptom and physiological testing data were collected concurrently. DNA methylation (DNAm) profiles were analysed longitudinally and compared to pre-pandemic DNAm data from healthy controls.

**Results:** While peripheral blood mononuclear cells (PBMCs) showed no significant changes, longitudinal DNAm changes were observed in neutrophil- and macrophage-enriched fractions. The changes were significantly associated with symptoms and physiological measures. Pathway analysis showed enrichment for cellular processes involved in cardiac function.

**Conclusions:** We identified longitudinal DNAm changes in lung immune cells associated with pathways linked to cardiac function. These changes correlate with symptom burden and heart and lung metrics. The results suggest potential disease mechanisms and aid the development of diagnostic tools.

## Introduction

Over 776 million COVID-19 cases have been reported globally, with around 65 million individuals experiencing persistent symptoms lasting months or even years after the initial infection. This condition, known as post-COVID-19 condition (PCC), is defined by the World Health Organization as new or ongoing symptoms at least three months after a COVID-19 infection, persisting for at least two months and not explained by other diseases (1–3). Patients with PCC exhibit a wide range of symptoms affecting multiple systems, including the cardiovascular, neurological, cognitive, and respiratory systems (4,5).

Several hypotheses have been proposed to explain the underlying mechanisms of PCC, such as viral persistence, latent virus reactivation, autoimmunity, neutrophil dysregulation, and inflammation-triggered changes (3). Despite significant progress in characterizing the various manifestations of PCC, the biological mechanisms driving the diverse disease trajectories remain elusive (6).

The persistent and debilitating symptoms reported by PCC patients underscore the urgent need to unravel the molecular mechanisms underlying these effects. In recent years, several studies have shown how infections impact the epigenomic landscape of immune cells, but data on PCC remains exiguous (7–9). Our group recently studied non-hospitalized PCC patients and found a distinct DNA methylation (DNAm) pattern in peripheral blood mononuclear cells (PBMCs) compared to a healthy control group and individuals fully recovered from COVID-19 (10). Additionally, studies on initially severely infected hospitalized patients indicate persistent DNAm changes in blood immune cells even 3-12 months after COVID-19 infection (11,12). Recent research on blood methylation has revealed significantly affected transcription factors related to circadian rhythm (13). Mapping these DNAm changes to cellular pathways has shown alterations in angiotensin II, muscarinic receptors, histamine signaling pathways, mast cell degranulation, and pathways involving the innate viral response and systemic autoimmune diseases (10–12). However, the dynamics of epigenetic regulation can vary across different cell types, and while COVID-19 is primarily a pulmonary infection, the DNAm patterns of lung immune cells remain uninvestigated.

We aimed to identify DNAm changes in lung and blood immune cells of PCC patients, comparing them to pre-pandemic healthy control subjects at two time points, to assess the persistence of these changes over time. Additionally, we aimed to investigate the association between DNAm changes, symptoms, and cardiopulmonary abnormalities.

## Materials and methods

### Study cohort and clinical measurements

Patients over 18 years with persisting symptoms following COVID-19 and referred to the Department of Clinical Physiology at Linköping University Hospital for lung function and exercise capacity evaluation were invited to participate in the study. Inclusion criteria were symptoms equivalent to a clinical diagnosis of PCC. Exclusion criteria included comorbidities with symptoms from the heart and lungs, inflammatory disease or difficulty in understanding the Swedish language. During their initial visit at inclusion (T1), those who gave their written, informed consent completed further testing, including questionnaires, blood and induced sputum sample collection, and an invitation to return for a follow-up after one year with repeated testing. The control samples used for comparison were derived from healthy subjects in two earlier studies (14,15).

#### Questionnaire and physiology data

Every participant was inquired about their immunization status, the highest level of COVID-19 care received (hospitalization and emergency room/primary care visits), date of infection and first symptoms, current symptoms, and history of polymerase chain reaction (PCR) and/or antigen testing for COVID-19 status (Table 1). The participants answered validated questionnaires about their degree of fatigue (measured by the modified Post-Covid Functional Status questionnaire (16)), quality of life (EQ-5D questionnaire (17)) together with a visual analogue scale (VAS) scale of grading their overall health status, and dyspnea (the King’s Brief Interstitial Lung Disease (K-BILD) questionnaire (18) and the modified Medical Research Council (mMRC) questionnaire) (19).

**Table 1.** Demographic characteristics of 13 participants with PCC.

| Parameter | At inclusion (T1) | One year follow-up (T2) |
| --- | --- | --- |
| Female, n (%) | 10 (77%) |  |
| Age, mean ± SD | 50.5 ± 12.3 (27-74) | |
| BMI, mean ± SD | 26.9 ± 5.8 (19.2-40.0) | 27.2 ± 5.7 (18.9-39.8)* |
| Former/active smoker, n (%) | 3 (23%) / 0 |  |
| Education, more than 3 years at university level, n (%) | 6 (46%) |  |
| **Vaccinated with COVID-19 vaccine (1 shot or more)\*, n (%)** | 7 (54%) | 11 (85%) |
| **Positive SARS-CoV-2 spike/both spike and nucleocapsid\*\* antibodies in plasma, n (%)** | 12 (92%) / 2** (15%) | 12 (92%) / 2** (15%) |
| **Hospitalized for COVID-19, n (%)** | 0 (0%) |  |
| **Days since COVID-19 infection at inclusion, mean ± SD** | 328 ± 149 (139-492) | |
| **Visited the emergency department for COVID-19 infection, n (%)** | 5 (39%) |  |
| **Visited primary care for COVID-19/PCC related symptoms, n (%)** | 12 (92%) | N/A |
| **Current sick leave ≥50%, n (%)** | 0 (0%) | 4 (31%) |

\* Comirnaty (Pfizer/BioNTech) or Spikevax (Moderna);
\*\* Positive for spike and nucleocapsid antibodies.
BMI, body mass index; SD, standard deviation.

The participants underwent a symptom-limited, graded maximal cardiopulmonary exercise test (CPET) on a bicycle ergometer (eBike Basic, GE Medical Systems, GmbH, Berlin, Germany) with continuous measurement of breathing gases and ventilation (Vyntus CPX, Viasys Healthcare). Peak oxygen uptake was defined as the mean of the two highest consecutive 10-second averaged values at the end of the exercise and compared to predicted values to determine exercise capacity (20). Each participant also underwent lung function testing, including dynamic spirometry (Jaeger MasterScreen PFT, Carefusion, Hoechberg, Germany) performed before and 15 minutes after bronchodilation using 400 μg of salbutamol with subjects in the sitting position and wearing a nose clip. Forced expiratory volume in one second (FEV1) and forced vital capacity (FVC) were obtained according to current standards (21). Diffusion capacity for carbon monoxide was assessed using a single-breath carbon monoxide diffusion test (Jaeger MasterScreen PFT) according to American Thoracic Society and European Respiratory Society standards. Patients were seated and wore a nose clip during testing. Reference values by Hedenstrom et al. were used for the calculation of % of predicted values (22,23).

#### Symptom-physiology score

Symptom data from the questionnaires EQ5D (VAS score), mMRC and K-BILD (total score) together with physiology data from CPET (including peak oxygen uptake, ventilatory efficiency and breathing pattern (24)), lung function testing - dynamic spirometry (including diffusion capacity, FEV1 and FEV1 divided by FVC (FEV%)), were used together for a combined symptom-physiology score. A summary of the data and the cut-offs is outlined in Additional file 1: Table S1 and Table S2.

### Laboratory procedures

#### Lung immune cells

Sputum samples were collected and processed as previously described by Pehrson et al. (14), with the following modifications: premedication with an inhalation of the beta agonist salbutamol was administered during lung function testing within an hour before sputum induction. Consequently, no inhalation was given during the sputum sample collection procedure. The subsequent analysis of cells incubated with Dynabeads® coupled with anti-CD3 antibodies (to collect cells associating with CD3-positive cells) revealed they were enriched in neutrophils, hereafter referred to as “neutrophil-enriched fraction”). In contrast, the unbound cell fraction incubated with Dynabeads® coupled to anti-HLA-DR/human MHC class II antibodies was enriched in macrophages, hence referred to as “macrophage-enriched fraction”). All magnetic beads and antibodies were from Invitrogen, Thermo Fisher Scientific, Massachusetts, USA.

#### Blood collection and processing of PBMC and plasma

Immune cells from peripheral blood mononuclear cells (PBMC) were collected and processed as previously described (10,15) with the following modifications: Blood samples of 30 ml were obtained, plasma separation was performed on 15 ml of whole blood by centrifugation (2000 G, 15 min, 4°C), no cells were cryopreserved, instead aliquots were stored in –80°C before further processing. DNA extraction and quantification was performed as previously described (10).

#### DNA methylation (DNAm) analysis

Genome-wide DNAm data was generated using 250 ng of purified DNA with the Illumina Infinium Methylation EPIC 850K BeadChip array (Illumina Inc, San Diego, USA) by the core facility for Bioinformatics and Expression Analysis at Karolinska Institute, Stockholm, Sweden.

### Data analysis

#### Clinical parameters

The Pearson Chi-square (χ2) and Fisher’s exact test (if the number of observations was smaller than five) were used for categorical variables to assess differences in the proportion of patients with abnormal values in physiological and questionnaire data at T1 and T2. For the CPET and lung function parameters, the significance test compares the number of patients with abnormal values between T1 and T2, and for the questionnaire data, the significance test compares the average mean values between T1 and T2. Continuous variables were compared using an unpaired two-tailed t-test with a significance level of p < 0.05.

#### Preprocessing of DNAm data

The resulting raw intensity (IDAT) files containing the DNAm profiles for each cell type were analysed in R (v. 4.0.2) using the minfi package (v. 1.36.0). The data was pre-processed in several steps, including the removal of probes that failed in one or more samples, cross-reactive probes and probes including CpG sites in the X and Y chromosomes or containing known single nucleotide polymorphisms (SNPs). The sex chromosomes were removed from our data set as female X-inactivation skews the distribution of beta (β) values. After filtering, quality control was performed, and normalisation of the data was done with the subset-quantile within array normalisation method (SWAN), which corrects for any technical variation in the data and makes the DNAm data comparable across samples. To determine the proportion of methylation at each CpG site, arrays measure both the methylated (M) and unmethylated (U) signals. The β values (M/ (M+ U +100) and M values (log2(M/U)) were calculated for each probe per sample. The quality of the data was assessed before and after the normalisation. To identify underlying components of variation within the normalised data set, we performed singular value decomposition (SVD) analysis using the ChAMP package (version 2.19.3).

The cell type proportions in the samples were inferred using the Epigenetic Dissection of Intra-Sample-Heterogeneity (EpiDISH) package (version 2.15.1), a reference-based cell type deconvolution method for DNAm data using robust partial correlations for inference, based on the Houseman algorithm (25,26).

#### Differential DNAm analysis

To visualize similarities and differences between samples, a principal component analysis (PCA) plot was used. For comparison, control samples were obtained from previously published datasets generated using the Illumina Infinium 450K array (14). The datasets from the current study (EPIC array) and the control samples (450K array) were independently preprocessed and normalized. Subsequently, the normalized datasets were merged to include only the common CpG sites across all samples. Due to the control and case samples being processed in separate experimental batches, batch effect correction methods such as ComBat were not applied to maintain the integrity of the biological signals in the data. Longitudinal patterns of DNAm within lung and blood immune cells were analyzed, using the limma package in R (v. 3.54.2) (27) to identify CpGs that were differentially methylated between two time points. To fit a linear model to the data, blocking by patient study ID was used to account for random effects with cell types as covariates in the model. In addition, the analysis was adjusted for identified sources of variation that remained significant in the SVD analysis (sex, age, body mass index, and smoking) by incorporating them as covariates in the model. CpGs were classified as differentially methylated (DMCs) when the absolute mean methylation difference (MMD) between T1 and T2 measurement was >0.15 and Benjamini-Hochberg FDR-adjusted *p*-value (hereafter referred to as *q*-value) was <0.05. β-values of identified DMCs were visualized using a heatmap (ComplexHeatmap package in R, v. 2.14.0) (28). In addition to β-values from the 26 samples of the 13 participants with PCC, β-values from 6 control samples were included as a reference (14).

After the detection of DMCs between samples, the DMCs were mapped to their corresponding differentially methylated genes (DMGs) using the getFlatAnnotation function from the missMethyl package in R (v. 1.32.1) (29). A gene was considered as differentially methylated if it contained at least one DMC (Additional file 2: Table S4 and Table S7).

#### Disease pathways analysis

The identified DMGs were analyzed for significant associations with disease pathways from the Kyoto Encyclopedia of Genes and Genomes (KEGG) database and functional properties from the Gene Ontology database using the clusterProfiler package in R (v. 4.6.2) (30). Additionally, protein-protein interactions were explored using the STRING database (https://string-db.org/) and the Biological General Repository for Interaction Datasets (BioGRID) database (https://thebiogrid.org/).

## Results

DNAm data for 18 participants was generated, but two subjects were excluded in the preprocessing step due to batch effects and three subjects were removed because of missing follow-up sampling. Consequently, the analysis focused on data from the remaining 13 patients who had longitudinal DNAm data from both time points.

Characteristics of the 13 study participants with PCC who attended both the inclusion (PCC T1) and one year follow-up (PCC T2) visits are shown in Table 1. A summary of the symptoms experienced by the PCC-group is stated in Additional file 1: Table S3.

### Different DNAm patterns in patients versus controls in both the neutrophil- and macrophage-enriched fractions

First, we assessed the source of variance using SVD analysis of DNAm data of lung- and blood-derived samples. The analysis revealed the timepoint (T1 vs T2) to be contributing to the variance both in the neutrophil- and macrophage-enriched fractions, but not in PBMC (Fig. 1: left column), which was confirmed with the PCA analysis (Fig. 1: middle column) with clearly separated clusters. To estimate the cell profiles of the different samples, we performed EpiDISH analysis showing the levels of cell types in each sample (Fig. 1: right column). The neutrophil-enriched fraction showed high proportions of neutrophils, with significant differences between controls, PCC T1 and T2 as indicated (Fig. 1: right column). The macrophage-enriched fraction showed high proportions of monocytes (as expected, due to the shared lineage of monocytes/macrophages with similar DNAm patterns) and varying levels of neutrophils in controls, PCC T1 and T2. In the PBMCs all groups displayed normal cell type proportions, except neutrophils, which were elevated in PCC T1 compared to controls.

**Fig 1.**
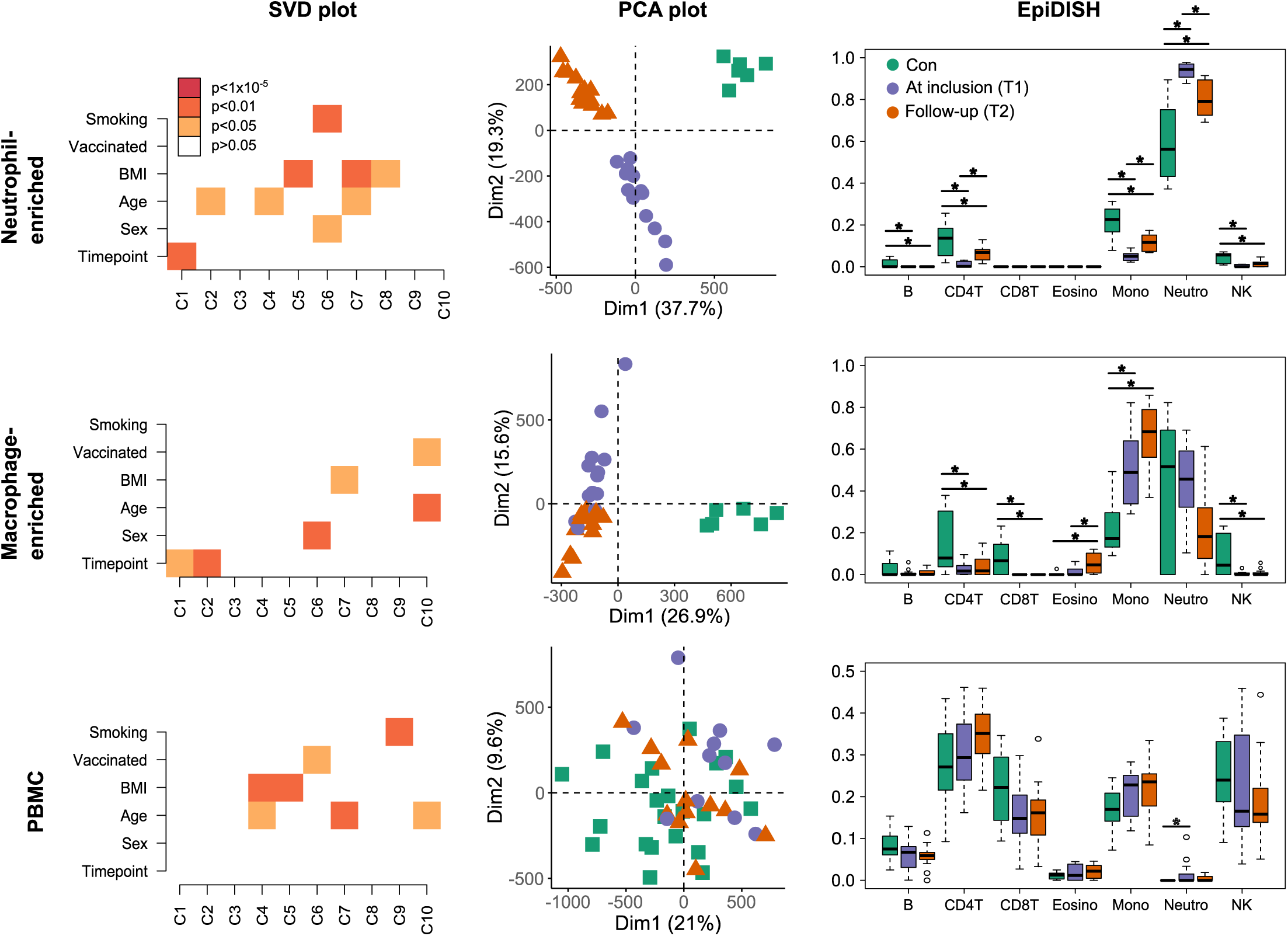
DNA methylation changes compared to controls in neutrophil-enriched fraction, macrophage-enriched fraction and PBMC. Singular value decomposition (SVD) analysis (left column). Values p>0.05 are considered non-significant (white areas on the plot). The x-axis shows components, and the y-axis shows demographic variables. Principle component analysis (middle column). The PCA plots showing the first two dimensions with explained percentage variance in brackets. Controls (Con), At inclusion for PCC (PCC T1) and follow-up for PCC (PCC T2). Cell type deconvolution analysis using EpiDISH (right column). X-axis showing cell types including B-cells (B), CD4-positive T-cells (CD4T), CD8-positive T-cells (CD8T), eosinophils (Eosino), monocytes (Mono), neutrophils (Neutro) and natural killer T-cells (NK). Y-axis showing estimated cell-type fraction. Statistical differences between groups was calculated with ANOVA, where significant differences (p<0.05) between the groups are shown with *.

### Longitudinal changes of DNAm patterns in the neutrophil- and macrophage-enriched fractions of lung cells

When comparing PCC T1 vs T2 longitudinally in the neutrophil-enriched fraction, we identified 446 DMCs, 70 hypermethylated and 376 hypomethylated with an MMD cut-off of 0.15 and *q*-value < 0.05. The controls’ corresponding β values are shown for reference (Fig. 2a). We explored the potential association between identified DMCs and disease pathways by mapping each DMC to the corresponding DMGs (Additional file 2: Table S4). We retrieved 334 KEGG pathways and identified the top 15 KEGG enrichment pathways with Benjamini-Hochberg *q*-value < 0.05, which were visualized (Fig. 2b). Among the top 15 pathways in the enrichment analysis, we observed cellular pathways connected to Wnt signalling and circadian entrainment as the most significantly enriched pathways. According to the BioGRID database, 87 DMGs interact with SARS-CoV-2 proteins (Additional File 1: Table S5).

**Fig 2.**
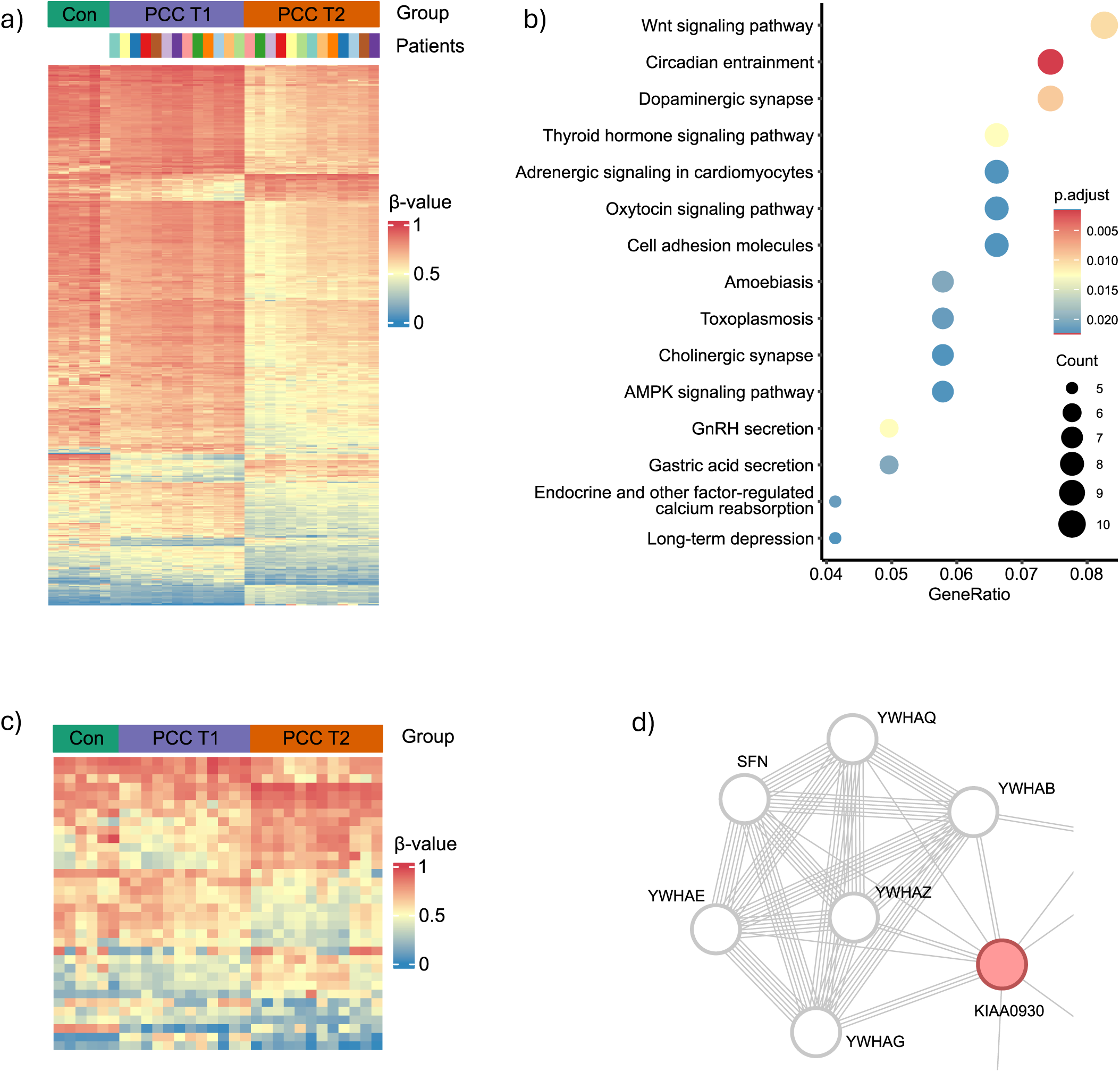
Longitudinal comparison of neutrophil- and macrophage enriched fraction. **a)** Heatmap for Controls (Con), PCC T1 (At inclusion) and PCC T2 (follow-up) showing the top 446 differentially methylated CpG-sites (DMCs) emerging from the comparison of PCC T1 with PCC T2 (MMD>0.15, FDR-corrected p-value <0.05) in the neutrophil-enriched fraction. **b)** KEGG pathway analysis for all DMCs, here showing the top 15 significantly affected gene pathways. P-value range (p.adjust) with attached colour. Count of affected genes in each pathway shown by size of the circle (Count). **c)** Heat map for Controls (Con), PCC T1 (At inclusion) and PCC T2 (follow-up) showing the top 34 differentially methylated CpG-sites (DMCs) emerging from the comparison of PCC T1 with PCC T2 (MMD>0.15, FDR-corrected p-value <0.05) in the macrophage-enriched fraction. **d)** STRING-db module with edges connecting KIAA0930 with a protein-protein interaction (PPI) network described as “SARS-CoV-2 targets host intracellular pathways and regulatory pathways” in Reactome pathways.

Next, we compared the DNAm data from the macrophage-enriched fractions at T1 and T2, identifying 34 common DMCs, 17 hypermethylated and 17 hypomethylated, when applying an MMD cut-off of 0.15 and *q*-value < 0.05 (Fig. 2c). The controls’ β values are shown for reference (Fig. 2c). In contrast to the neutrophil-enriched fraction, the DMGs corresponding to the identified DMCs (Additional file 2: Table S7) were not significantly connected to any KEGG pathways. In an overlap analysis between the DMGs identified in both of the lung-derived cell fractions (Additional file 1: Table S5 and Table S8), we identified one common DMG, KIAA0930, which has been implicated in hypoxia adaptation (31) and found differentially methylated after major surgery (32). STRING-db analysis of modules related to KIAA0930 revealed the protein as a node with multiple edges linking to proteins of the 14-3-3 protein family (Fig. 2d) (link to STRING-db analysis), which in Reactome Pathways is described as part of the “SARS-CoV-2 targets host intracellular pathways and regulatory pathways” (R-HSA-9755779).

### Connecting DNAm with a symptom-physiology score

The symptom-physiology score derived from self-evaluation questionnaires and physiology data at T1 and T2 highlight the symptom heterogeneity of the 13 participants, shown with individually coloured circles for each participant (Fig. 3a). Longitudinal symptom improvement was observed in seven patients, whereas three patients showed similar score both time points and three patients had worsening of the score at follow-up. We found 70 DMCs whose change in β value between T1 and T2 correlated (with a p-value <0.05) with changes in symptom-physiology score (Fig. 3b) using the method proposed in (33). These 70 DMCs were mapped to 54 unique DMGs (Additional file 2: Table S6). STRING-db analysis of these DMGs revealed a connection to pathways associated with cardiac function (Fig. 3c), (link to the STRING-db analysis). One of the identified modules of connected DMGs involved proteins linked to the KEGG pathway “viral myocarditis” (hsa05416) (Fig. 3d) (34).

**Fig 3.**
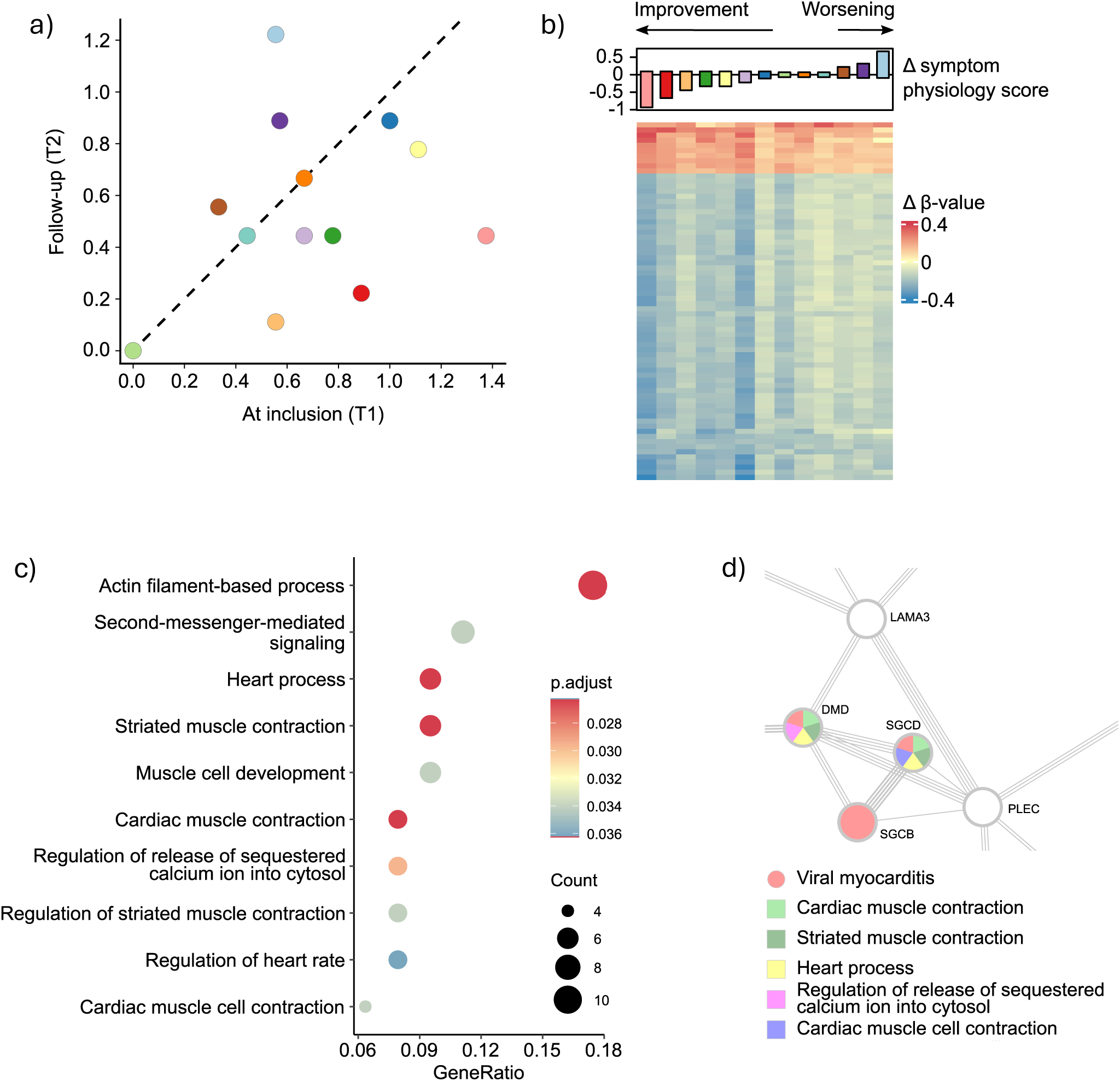
Symptom-physiology score. **a)** Symptom-physiology score plot with means for all individuals at inclusion (T1)/follow-up (T2). Dashed line separates participants with higher score at T2 compared to T1 to the left of the line from lower score at T2 to the right. **b)** Heat map with delta β value of 70 DMCs that correlate with delta symptom-physiology score. Delta symptom-physiology score is calculated with the score value at T2 subtracted to the value at T1, same method for delta β value. **c)** Gene Ontology enrichment analysis for DMCs that correlate with delta symptom-physiology score, here showing the top 10 significantly affected pathways. P-value range (p.adjust) with attached colour. Count of affected genes in each pathway shown by size of the circle (Count). **d)** STRING-db module describing a PPI network connecting proteins involved in cardiac function (Gene Ontology, marked with squares in labels) and viral myocarditis (KEGG pathways, marked with circle in label).

## Discussion

This pilot study of 13 non-hospitalized patients with varying severity of PCC is the first to provide longitudinal DNAm data from lung-derived immune cells. We identified distinct DNAm patterns in the lung immune cells with longitudinal changes observed in neutrophil- and macrophage-enriched fractions, but not in blood (PBMC). Additionally, we determined a significant association between COVID-19-induced DNAm changes and the symptom-physiology score, which mapped to biological pathways related to cardiac function. Interestingly, cardiac-related complications, such as dysautonomia and exercise intolerance, are often the most common symptoms in PCC, yet no clear pathophysiological explanation is currently available (5,35,36).

The presence of DMGs linked to genes involved in cardiac function in DNA from lung immune cells remains unclear. One possible explanation for such ubiquitously observed modifications to the epigenome could be that alterations in DNAm patterns are induced either by the host as a defence mechanism, by the virus to manipulate host defences, or a combination of both, in many cell types in the body.

Our identification of DMGs that correlate with the symptom-physiology score may imply that epigenetic alterations are involved in the pathophysiology underlying the symptoms experienced by the patient. From a clinical-diagnostic perspective, these results are relevant for further development of diagnostic tools and a better understanding of disease mechanisms.

The method we used to purify cells from the lungs yielded mixed cell fractions rather than highly homogeneous isolates, making it challenging to definitively attribute the observed DNAm changes to specific cell types. The level of neutrophils in the neutrophil-enriched fraction was higher in T1 than in T2 and controls. Although cell type proportions were included as covariates in our analysis to mitigate their confounding effects, this cell type may still substantially contribute to the observed DNAm changes emerging from the longitudinal analysis. At T1, a significant proportion of neutrophils contaminated the macrophage-enriched fraction, and this problem was less obvious at T2. One reason for this result may be that patients with PCC have a higher level of circulating neutrophils in blood also affecting the lungs. At T1, we found slightly but statistically significantly higher levels of neutrophils in the PBMC fraction, which normally should not contain neutrophils. One possible explanation is that the gradient centrifugation steps used to enrich PBMCs exclude neutrophils due to their higher granularity (=higher density). In support of this observation, several studies have identified a higher level of immature neutrophils after COVID-19 infection (37,38). In one study, the authors were able to correlate the proportion of circulating immature neutrophils to COVID-19 disease severity (37). In another study, low-density neutrophils found in the PBMC fraction were linked to COVID-19-induced, persistent pulmonary dysfunction and Neutrophil Extracellular Trap (NET) formation (38). Furthermore, other studies have seen more immature progenitor cells in blood of patients after severe COVID-19, pointing towards epigenetic reprogramming in the progenitor cells in the bone marrow as a mechanism for affected immune function (39). Further studies are needed to characterize the neutrophil phenotype observed in PCC patients and their possible contribution to the disease.

We identified modifications to the DNA methylome that map to intracellular pathways crucial for cellular defence mechanisms and endocrine functions. Among the top 15 enriched KEGG pathways from DMGs in the T1 vs T2 comparison are circadian entrainment, dopaminergic synapse, adenosine monophosphate-activated protein kinase (AMPK) signalling pathway, and long-term depression, all of which are clinically relevant due to their connection to symptoms in PCC. This finding aligns with recent studies: one showing that metformin, an AMPK activator, reduces the incidence of PCC (40), and the one indicating significant effects on the methylation levels of transcription factors for circadian rhythm in blood (13).

To work around the limitation with variable cell type proportions in the lung fractions, we looked for DMGs present irrespectively of the cell-enrichment method. We found a common gene in both the neutrophil and macrophage-enriched fraction, KIAA0930, which is proposed to be important in lung cancer development (41). Protein interaction analyses provide further insights into the possible functional roles of KIAA0930. In the STRING-db protein network, KIAA0930 is shown to interact with six proteins involved in the SARS-CoV-2 pathway in the Reactome database, while BioGRID database identifies a direct interaction between KIAA0930 and ORF3A (42), a SARS-CoV-2 accessory protein known to modulate immune responses and cellular stress pathways. These interactions suggest that KIAA0930 is at the crossroads of host-virus dynamics in COVID-19, positioning it as a potential link to post-COVID-19 pathophysiology.

In conclusion, we provide evidence for longitudinal changes in the epigenome of lung immune cells from patients with PCC, which correlate with changes in symptoms and physiological measures over time. Given our limited sample size, larger cohorts are needed to clarify the relationship between DNAm patterns and specific cell types. Our findings propose a potential role of DNAm alterations in explaining symptoms associated with PCC.

## Acknowledgements

We express our greatest thanks to the Core facility at Novum, Bioinformatics and Expression Analysis, at the Karolinska Institute, Stockholm, Sweden and the staff at the Department of Clinical Physiology, Linköping University Hospital, Linköping, Sweden.

Thanks to all the participants for the help and dedication in the study. We are thankful for the help by Lovisa Karlsson with the laboratory analyses, and for the support by Helena Engström at the Department of Pulmonary Medicine. We also thank Alexander Vergara for his help with bioinformatics analysis.

The data handling was enabled by resources provided by the National Academic Infrastructure for Supercomputing in Sweden (NAISS) at the National Supercomputing Centre (NSC), Linköping University.

This work was supported by the Swedish Heart Lung foundation, as a sub-project within the project “Deciphering infection related epigenetic dynamics to explain the variability of human susceptibility to COVID-19 and tuberculosis” (Grant no. 2022-0034). Funding was also granted from local grant (Student to Docent) and regional ALF-grant in Region Östergötland, Sweden. The enabling of data handling was partially funded by the Swedish Research Council (Grant no. 2018-05973).

## Ethical approval

The study was approved in April 2021 by the Swedish Ethical Review Authority (Dnr. 2021-01620). The participants provided their written informed consent to participate in this study.

## Conflict of interest

ML, MG, SS and DM are founders of a company, which has the ambition to develop DNAm-based biomarkers for a variety of conditions, including PCC. The other authors declare no competing interests.

## Author contributions

FN, KH and ML designed the study. KH and ML secured project funding. FN and KH coordinated the study design and inclusion. FN collected the blood and sputum samples. FN conducted the laboratory analyses. JS and SS performed the data curation, bioinformatics analysis and interpretation. JS, FN and SS performed the statistical analysis. JS prepared the figures and tables. SS, DM, MG, MR and ML evaluated the results. JS, FN and SS drafted the manuscript. All authors were involved in reading and revising the manuscript prior to submission, and all approved the submitted version.

## Data sharing and availability

The deidentified processed data that support the findings of this study are available from the corresponding author upon reasonable request.

## Additional files

Additional files provided with this manuscript:

**Additional File 1** (docx): Contains Tables S1-S3, S5, and S8.

- Includes data from the symptom-physiology score, with a summary of statistics and cut-offs.
- Provides a table summarizing symptoms in the PCC group.
- Lists DMGs found in neutrophil- and macrophage-enriched fractions.

**Additional File 2** (xlsx): Contains Tables S4, S6, and S7.

Detailed longitudinal analyses of DMGs from neutrophil- and macrophage-enriched fractions and the symptom-physiology score.

**Figures** (pptx): Includes Figures S1-S3 with corresponding figure legends.

## Notes

### Author Declarations

Ethics committee of Swedish Ethical Review Authority (Dnr. 2021‐01620) gave ethical approval for this work.

