## Additional file 1 for "Longitudinal epigenetic rewiring in lung immune cells in patients with post-COVID-19 condition"

### Supplementary tables (S1-S3, S5, S8)

**Table S1 – Symptom physiology score – cut offs for the different scoring points.**

| Scoring | Ventilatory efficiency (VE/VCO2 -slope) | Peak oxygen uptake (%) | Breathing pattern | Forced expiratory volume in 1 second (FEV1) (%) | FEV1/forced vital capacity (FVC) (%) | Diffusion capacity (%) | modified Medical Research Council questionnaire (mMRC) | EuroQoL 5 Dimensions (EQ-5D) visual analogue scale (VAS) | King's Brief Interstitial Lung Disease (K-BILD) total score |
|---|---|---|---|---|---|---|---|---|---|
| 0 - normal | <30 | >90 | normal | >80 | >95 | >75 | 0-1 | 75-100 | 75-100 |
| 1 - mildly abnormal | 30-34 | 80-89 | mildly abnormal | 70-79 | 85-94 | 50-75 | 2 | 50-74 | 50-74 |
| 2 - abnormal | 35-39 | 70-79 | abnormal | 50-69 | 75-84 | 30-49 | 3 | 25-49 | 25-49 |
| 3 - very abnormal | >40 | <70 | | 30-49 | 65-74 | <30 | 4 | 0-25 | 0-24 |

**Table S1. Symptom physiology score.** The absolute values for the nine parameters (columns), were translated to a comparable scoring between 0 (normal value) - 1 (mildly abnormal) - 2 (abnormal) - 3 (very abnormal) (rows). The values for each scoring point were set according to the reference limits in clinical practice for the different tests, respectively. For the physiology data from CPET, ventilatory efficiency was defined using the ventilatory equivalent of carbon dioxide (VE/VCO2), and the lowest (nadir) value >30 was considered abnormal and as inefficient ventilation. A peak oxygen uptake <90% of the predicted value was considered abnormal (1). The breathing pattern of each subject during exercise was categorized as normal, borderline, or abnormal, as previously described (2). The normal value of FEV%, FEV1 and VC was set to 75% and above (3). Cut off for EQ-5D VAS was set to 75 according to known normal means for different age groups (4,5). For mMRC normal test value was considered to be 0-1 (6). The K-BILD total score was considered abnormal with a value below 75. The comparable scoring points for the nine parameters were calculated as a mean, both at T1 and T2, to be compared individually within the PCC-group.

**Table S2 – Summary of data from the CPET, lung function testing and questionnaire data used in the symptom-physiology score.**

| Variables | Normal value | At inclusion (T1) | | Follow-up (T2) | | Significance (p<0.05) |
|---|---|---|---|---|---|---|
| | | Average mean value | Proportion of patients with abnormal value (percentage) | Average mean value | Proportion of patients with abnormal value | |
| **CPET PARAMETERS** | | | | | | |
| **Ventilatory efficiency (VE/VCO2-slope)** | < 30 | 30.4 | 54% (7/13) | 29.1 | 38% (5/13) | ns |
| **Peak oxygen uptake** | ≥ 80% of predicted | 97.0% | 23% (3/13) | 102.0% | 8% (1/13) | ns |
| **Breathing pattern** | | | 69% (9/13) | | 46% (6/13) | ns |
| **LUNG FUNCTION PARAMETERS** | | | | | | |
| **FEV1** | > 80% | 102.5 | 0% (0/10) | 105.4 | 0.0 (0/10) | ns |
| **FEV% (FEV1/FVC)** | > 95% | 106.8 | 0% (0/9) | 107.7 | 0.08 (0/9) | ns |
| **Diffusion capacity** | > 75% | 96.3 | 0% (0/9) | 88.3 | 0.23 (2/9) | ns |
| **QUESTIONNAIRE DATA** | | | | | | |
| **mMRC** | 0-1 = no significant dyspnea | 1.7 | 0.50 (6/12) | 1.9 | 0.75 (9/12) | ns |
| **EuroQoL 5 Dimensions (EQ5D)** | 0 = worst imaginable, 100 = best imaginable | 52.0 | | 56.8 | | ns |
| **King's Brief Interstitial Lung Disease (K-BILD) total score** | low number = worse | 73.6 | | 75.7 | | ns |

**Table S2.** For the CPET and lung function parameters, the significance test compares the number of patients with abnormal values between T1 and T2. For the questionnaire data, the significance test compares the average mean values between T1 and T2. Continuous variables were compared using an unpaired two-tailed t-test and categorical variables were examined using the Pearson χ2 test or Fisher's exact test (if the number of observations was smaller than five).

**Table S3 – Summary of the PCC symptoms. Data from the questionnaire at T2 from the 13 participants.**

| Category | Symptom | Number of subjects (percentage) |
|---|---|---|
| **Physiological** | Palpitations | 6 (46%) |
| | Impaired mobility | 2 (15%) |
| | Dry cough | 3 (23%) |
| | Chest pain/discomfort | 4 (31%) |
| | Joint/muscle pain | 7 (54%) |
| | Throat pain | 2 (15%) |
| **Neurological** | Loss of smell | 3 (23%) |
| | Loss of taste | 4 (31%) |
| | Dizziness | 4 (31%) |
| | Headache | 4 (31%) |
| | Fatigue | 10 (77%) |
| **Psychiatric** | Anxiety/depression | 4 (31%) |
| **Other** | Gastrointestinal problems | 2 (15%) |
| | Skin problems | 5 (38%) |

**Table S5 – List of DMGs in neutrophil-enriched fraction with SARS-CoV2 interactions in the BioGRID database.**

| | | | | | |
|---|---|---|---|---|---|
| PTPRF | TRAM2 | ACOT7 | RBM47 | ZCCHC14 | AGTPBP1 |
| INPP5A | NCOR2 | EHMT1 | TNRC6B | GSK3B | DICER1 |
| KCNE4 | EEF2 | MYO5A | MKLN1 | PRKAG2 | PAX7 |
| KIAA0930 | SH3BP4 | DNMBP | TACC2 | ZDHHC7 | SREBF1 |
| FARP2 | TJAP1 | TRAF7 | FRMD5 | CUEDC1 | PFKP |
| NOTCH4 | CELSR1 | GGT1 | PLEC | MCC | PLCB2 |
| POU6F2 | DNAJB1 | PTPRS | TSPAN14 | TAPBP | UPP1 |
| POLM | TNFAIP2 | COIL | AGO2 | GALNT2 | HNRNPR |
| CYP4V2 | SLCO4A1 | HCN2 | CSPG4 | TMPRSS4 | C7orf50 |
| EVA1B | CCDC50 | SPTBN2 | PRKCA | TRIM29 | SLC16A5 |

| | | | | | |
|---|---|---|---|---|---|
| FOXK1 | PLEKHA6 | CUX1 | CTNND1 | SYCE1 | SMARCA4 |
| MACROD1 | CNP | CDC73 | SUCLA2 | BNIP3 | TAF4 |
| PACSIN2 | PLD6 | SGCD | LAMB3 | BAIAP2 | SLC1A4 |
| LAMA3 | WDR46 | SMAD3 | CORO1C | CIT | MPZL2 |
| LRRK1 | SPEN | TSC2 | | | |

**Table S8 – List of DMGs in macrophage-enriched fraction with SARS-CoV2 interactions in the BioGRID database.**

| | | | | | |
|---|---|---|---|---|---|
| CORO7 | FFAR2 | NOTCH1 | EPB41L5 | USP10 | ENO1 |
| ASB1 | TFDP1 | EGLN1 | KIAA0930 | EIF4G1 | PACSIN1 |
